# Informal Sector Employment and the Health Outcomes of Older Workers in India

**DOI:** 10.1101/2022.03.24.22272875

**Authors:** Poulomi Chowdhury, Itismita Mohanty, Akansha Singh, Theo Niyonsenga

## Abstract

A large proportion of the older population in India constitutes an undeniable share of workforce after the retirement age. This stresses the need to understand the implications of working at older ages on health outcomes. The main objective of this study is to examine the variations in health outcomes by formal/informal sector of employment of older workers using the Longitudinal Ageing Study in India. Using binary logistic regression models with interaction terms, the results of this study affirm that type of work does play a significant role in determining health outcomes even after controlling socio-economic, demographic, life-style behaviour, and work characteristics. The risk of Poor Cognitive Functioning (PCF) is high among informal workers, while formal workers suffer greatly from Chronic Health Conditions (CHC) and Functional Limitations (FL). The presence of CHC among informal workers is more damaging because it elevates their FL, whereas for formal workers presence of FL is unfavourable because it deteriorates their cognitive functioning. Therefore, present study underscores the relevance of policies focusing on providing health and healthcare benefits by respective economic activity and socio-economic position of older workers.

## Introduction

Globally, ageing population has become a common phenomenon owing to reduction in fertility and mortality rates, and these integrated effects have altered the age-sex composition as well as labour force participation towards higher ages (1, 2). Speculation indicates that the world population aged 60+ will be around 2 billion in 2050 from 900 million in 2015 (3). By 2050, nearly 80 percent of the older population will reside in low-middle income countries (3). Ageing process in developed nations has slowed down the economic growth through insufficient labour force. These nations have implemented policies aiming to boost labour force market by motivating the older population to work past the age of 65 years (4-10). Consequently, in recent years, the share of older workers has amplified substantially in developed nations (4, 5, 7, 9-13).

Apart from these nations, the developing countries are also experiencing shifts in age-structures with a tremendous pace. United Nations (UN) report (2017), states that almost 60 percent of the older people are currently living in developing countries, and is growing rapidly compared to developed countries (14). Alas, the provision and implementation of pension benefits or retirement programs are less prevailing in developing nations. Apparently, only 20 percent of this population is entitled to any pension related benefits, yet most of them rely on family support system (15). Accordingly, significant proportion of older people are active in labour market in developing countries (2, 16-20) than developed countries (21).

India, the second most populous nation in the world, is also facing remarkable increase in ageing population, reportedly 8.6 percent (104 million) of the total population in 2011. This figure is expected to escalate to about 20 percent and, in terms of absolute numbers, the country will be a home for 319 million older individuals by 2050 (22). Soon this older population will surpass the young population below 14 years (23).

Ageing is normally associated with chronic health conditions which upsurge later in life (24-27). As evident from prior studies, more than half of the older people endure non-communicable diseases (NCDs), while one-fourth are affected by multimorbidity (28, 29). Across India, these chronic diseases exhibit huge heterogeneity in terms of socio-economic conditions, place of residence and gender (25, 26, 30). Further, estimated figures suggest that the burden of NCDs will constitute a large share of the national disability (31). Projected numbers demonstrate that roughly 45 percent of the health burden will be borne by older people (32, 33), making the health requirements of older people comparatively higher than other age groups.

Specific social security, poverty alleviation and social welfare programs have been launched to address the challenges associated with older population’s social and health conditions. Certain policies and programs implemented by the ministries, namely, National Policy on Older Persons, National Social Assistance Programme, National Policy for Senior Citizens, Indira Gandhi National Old Age Pension Scheme, and Mahatma Gandhi National Rural Employment Guarantee Scheme, have failed to offer adequate financial assistance to support the older persons’ requirements, particularly to those in unorganized working sector and below poverty line (34-36). Rashtriya Swasthya Bima Yojana was also introduced to provide health insurance to the workforce engaged in unorganized sector but unable to perform well because it failed to capture below poverty line families, tribal blocks, and impoverished sections of the society (37-39). To achieve the sustainable development goal of universal health coverage, the government of India has approved Ayushman Bharat Yojana (ABY) in March 2018. It is an ambitious scheme to provide financial health protection for 500 million Indian population belonging to vulnerable sections. However, like every health program in India, the success of ABY lie on overcoming existing issue like public and private sector governance, quality control, stewardship and health system organization (40).

Unfortunately, majority of the Indian older population are unable to access healthcare services after the retirement age (60 years and above) due to paucity and poor coverage of universal health and pension programmes. Therefore, to manage the livelihood and healthcare needs, older people are compelled to work after the retirement age (19, 41). Census of India (2011) figures illustrate that a large proportion (33 million) of older people are working after the retirement age, especially in informal sector. However, informal sector provides financial support to only marginal level of workforce after the retirement (20). Even in formal sector, nearly 10 percent of the population engaged in selected organized work places receive the benefit of social or voluntary health insurance schemes (42). The absence of proper financial and health care schemes can hamper the health conditions of older people indulged in economic activities. It makes older workforce as one of the most vulnerable groups in India.

Thus, given the rising magnitude of older workers, it becomes essential to understand the extent to which their health conditions are associated with the type of employment. However, no studies till date, nationally or internationally, have emphasised on this aspect. Keeping this in mind, the present research focused on health outcomes/conditions, specific to older workers who are engaged in formal and informal sector of employment. It hypothesized that people engaged in informal employment work will experience more unfavourable health outcomes, that is, high rates of chronic health conditions, functional limitations, and poor cognitive functioning, compared to those in formal sector of employment. Research findings will help in addressing the important policy issues considering the extent of variation in health among older workers in India (41, 43).

## Research Framework

Persisting health problems among older people constitutes a longstanding concern for researchers and policy makers as the prevalence of morbidity is relatively high in later ages. It is acknowledged that older people continue to participate in the labour force despite of the health risks and socio-economic challenges, particularly in developing nations (2, 19). Kalwij, Kapteyn (7) stated that health is multidimensional in nature and the effect of the work engagement on health varies with health indicators assessed. Evidently, prior studies have discovered that work engagement has a pronounced effect on physical (11, 12, 44-50) and mental health (10-12, 46-48, 50-52). Few studies asserted that engagement in low paid jobs negatively influences physical health of the older people (12, 44). Besides, working longer with degraded health conditions can have severe repercussions like need for long-term care, mental health issues and functional disability (11, 12). Nevertheless, prolonged older age work engagement has also a beneficial effect on mental health (10-12, 47, 50). This holds true in the study of Japan and Korea which describes that working in later life is generally associated with financial security and strong social network leading to better cognitive functioning and less depressive symptoms (11, 50).

The relationship between work engagement and health conditions may vary by type of occupation (51, 53, 54). This relationship is also shaped by various socio-economic and demographic attributes (2, 5, 12, 18-20, 44, 51, 52, 55), work characteristics (11, 12, 55-57) and lifestyle behaviours (11, 16, 57-59). Altogether, previous studies reflected a robust relationship between work engagement and health, and taking these into account, a research framework has been conceptualized. In addition, strong relationships within health indicators also have been described in previous studies. For instance, chronic health conditions (CHC) amplifies the functional limitations (16, 49), whereas functional limitations and CHC influences cognitive functioning of older people (11, 49, 51). Moreover, CHC may exert its impact on individual poor cognitive functioning either directly (direct effect) or through functional limitations (indirect effect). The research framework reflecting the relationships between type of work and health outcomes is depicted below (Fig 1), along with adjustment factors, including lifestyle behaviours, socio-economic and demographic characteristics as well as work characteristics.

**Fig 1:**
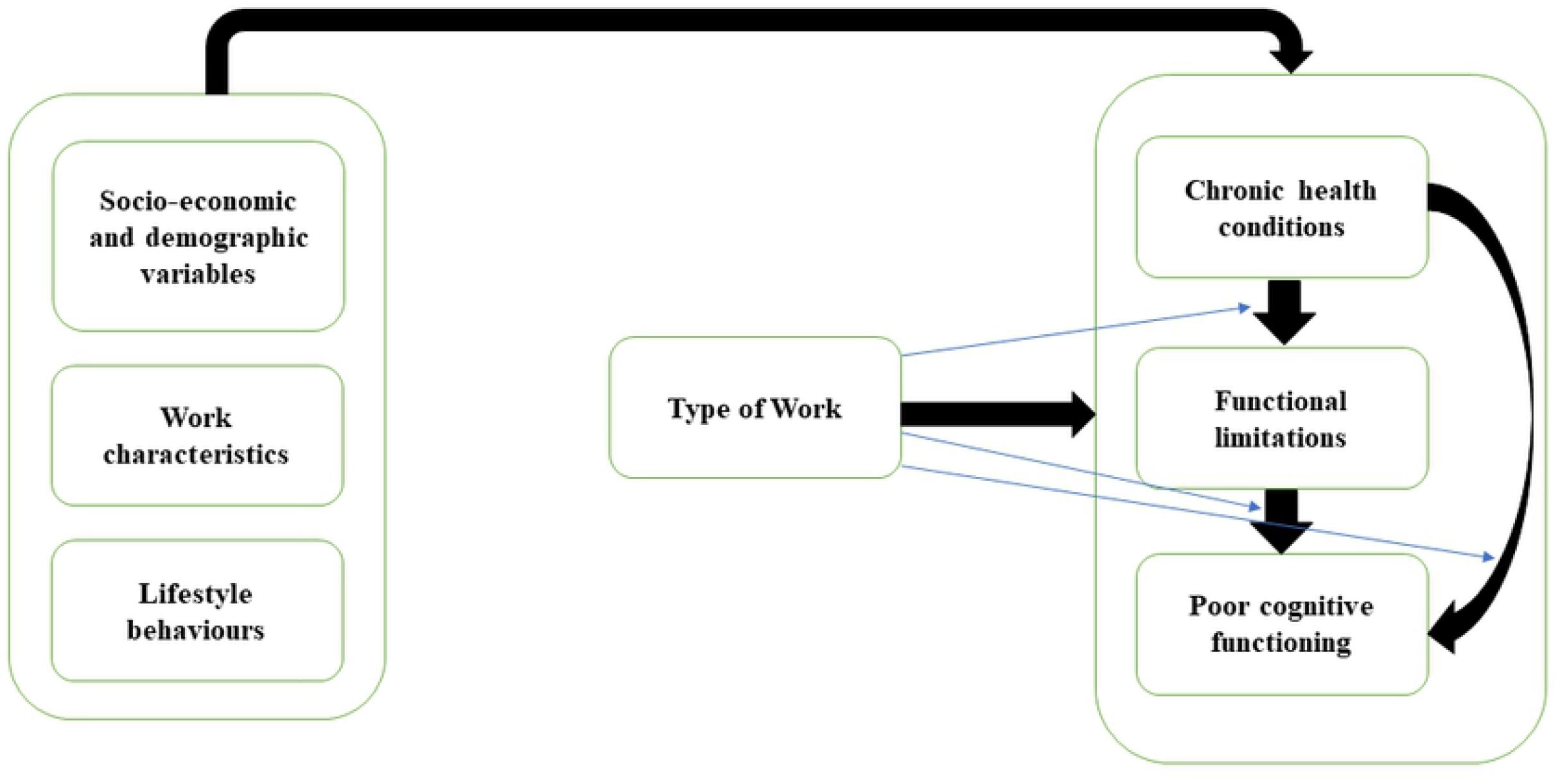
Research framework. Note: Blue lines represent moderation process.

## Materials and Methods

### Study design

The present study has employed nationally representative data of Longitudinal Ageing Study in India (LASI). It is based on a cross-sectional design as it uses only one wave (baseline) of the LASI data.

### Data source

The LASI baseline survey (2017-18) was conducted with the joint collaboration of International Institute for Population Sciences, Harvard TH Chan School of Public Health, and the University of Southern California (60). This LASI survey followed a multistage stratified cluster sample design and collected information on 72,250 Indian adults aged 45 years and above, across all states and union-territories. Out of which, the sample in this study contains 10,746 older people (60 years and above), currently engaged in the workforce.

The LASI survey aimed to collect the longitudinal data of older adults which include the information on social and economic wellbeing, burden of diseases, functional health and healthcare based on internationally comparable research design and tools. It created a foundation for reliable and acceptable data for national policy and long-term scientific research. Further, it provided in-depth information of economically active older population, workforce participation across older ages and different sectors, perceived economic security, work characteristics, vulnerability and expectations.

### Outcome variables

Following the research framework, the present study has emphasized on three health outcomes i.e., chronic health conditions (CHC), functional limitations (FL), and poor cognitive functioning (PCF). Details of these health outcomes are given below:

#### 1. Chronic Health Conditions

The CHC is summarized using nine self-reported health conditions. These health conditions are hypertension, diabetes, cancer, chronic lung disease, chronic heart diseases, stroke, arthritis, neurological problems, and high cholesterol. The format used in the questionnaire is “Has any health professional ever diagnosed you with any of the chronic conditions or diseases…..?”. Based on this information, CHC is categorized into 0 (none) and 1 (at least one health condition), where 0 labeled as ‘No’ and 1 as ‘Yes’.

#### 2. Functional Limitations

The FL is constructed using 13 everyday activities which are generally termed as activities of daily living (ADL) and instrumental activities of daily living (IADL). Out of the 13 activities, the first 6 are related to ADL, while rest are associated with IADL. The question format of FL is “Because of health or memory problem, do you have any difficulty with any of the activities….?”. The FL is dichotomized as 0 (no limitations) and 1 (at least one limitation), where 0 labeled as ‘No’ and 1 as ‘Yes’.

#### 3. Poor Cognitive Functioning

For the assessment of poor cognitive functioning health outcome, the present research has followed the LASI report which uses cognitive module of Health and Retirement Study (HRS) involving memory (0-20), orientation (0-8), retrieval fluency (0-61), arithmetic function (0-9), executive function (0-4), and object naming domains (0-2). First these indicators are normalized by employing following formula:

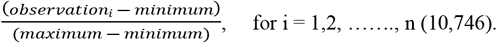

This normalization helps in rescaling the indicators between 0 to 1 (61). Then, a principal component analysis (PCA) is applied to create a composite score of cognitive functioning, where minimum score represents poor cognitive while higher score reflects better cognitive functioning (62, 63). This score is further divided into three equal parts (tertile groups), where first tertile is coded as 1 (to represent poor cognitive) while rest are coded as 0 (otherwise).

### Type of work

LASI survey follows International Classification of Occupation 2015 to categorize the occupation types. These categories are further grouped into formal and informal work based on the guidelines provided by 66^th^ round of National Sample Survey Organization report which adopts the National Classification of Occupation 2004 (64, 65). The type of work variable is dichotomized as 0 (formal) and 1 (informal).

### Other independent variables

As per the research framework, three main dimensions of covariates are considered, that is, socio-economic and demographic, work characteristics, and life-style behavior. Socio-economic and demographic dimension includes gender (male, female), age groups (60-65, 65+), caste groups-based on the access to wealth, power and privilege (general, Scheduled Tribe (ST), Scheduled Caste (SC), Other Backward Class (OBC)), religion (Hindu, Muslim, others), educational level (low, middle, high), marital status (currently married, others), place of residence (rural, urban), wealth (low, medium, high), and household size (1, 2, 3, 4+). Work characteristics include working hours per week (less than 24 hours, 24-48 hours, 48+ hours) and monthly wages. The life-style behavior variables are Drinking alcohol (no, yes), smoking/consuming tobacco (no, yes), physical activities consisting vigorous activities (never, rare, everyday), moderate activities (never, rare, everyday), and yoga/pranayam (never, rare, everyday). Apart from these variables, the region is also taken as independent covariate which involves North (Jammu & Kashmir, Himachal Pradesh, Punjab, Uttarakhand, Haryana, Delhi, Rajasthan), Central (Uttar Pradesh, Chhattisgarh, Madhya Pradesh), East (Bihar, West Bengal, Jharkhand, Odisha), North-East (Arunachal Pradesh, Nagaland, Manipur, Mizoram, Tripura, Meghalaya, Assam), West (Gujarat, Maharashtra, Goa), South (Andhra Pradesh, Karnataka, Kerala, Tamil Nadu, Telangana), and Union territories (Chandigarh, Daman & Diu, Dadar & Nagar Haveli, Lakshadweep, Puducherry, Andaman & Nicobar).

### Statistical Analysis

Descriptive summary statistics were compiled using proportions and means with associated standard deviation. The study utilized bivariate analysis with chi-square test of association and multivariate analysis following the research framework to investigate the relationships depicted in Fig 1.

First the percentage of formal and informal older workers aged 60 year and above are calculated by socio-economic and demographic variables. Then, the prevalence rate of CHC, FL and PCF are calculated by type of work and chi-square test is applied to measure the significance of association. Lastly, sequential multiple logistic regression models are employed as the outcome variables are dichotomous. In the first model (Model-1), chronic health conditions (CHC) outcome is the function of type of work only, while in the second and third models, the effect of type of work on CHC is assessed by controlling for socio-economic and demographic variables (Model-2), and work characteristics, life-style behavior and regions (Model-3) respectively. For functional limitation (FL) outcome, Model-1 includes the type of work and CHC, while in Model-2, the interaction term CHC*Type of work is added along with socio-economic and demographic indicators. Similarly, Model-3 adjusts for work and life-style characteristics and regions, as in the case of CHC. For poor cognitive functioning (PCF) outcome, Model-1 includes the type of work, CHC and FL. The interaction terms CHC*Type of work and FL*Type of work, along with socio-economic and demographic indicators, are added in its second model (Model-2). Finally, in the third model (Model-3), the effect of type of work on health outcomes is observed by controlling all variables associated with socio-economic and demographic, work characteristics, life-style behavior dimensions as well as the respective interaction terms. Below is the description of all models by health outcomes (Table 1), following the research framework (Fig 1).

**Table-1:**
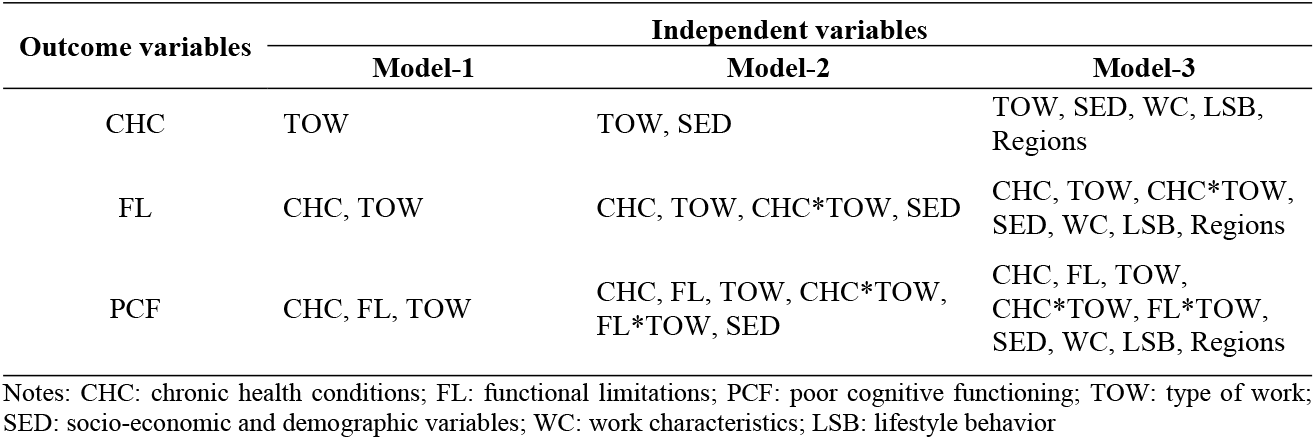
Description of logistic regression models.

## Results

### Descriptive analysis results

Table 2 shows that approximately one-third of the older population is currently working, out of which 73.1 percent are engaged in informal employment activities. Predominantly, the percentage of older workforce is highest among males, Hindus, OBC group and in rural areas. More than three-fourth of working population belongs to lower education level, while one-third of them lies in low wealth status. Drinking alcohol and smoking/consuming tobacco among older workforce is 13.1 percent and 50.8 percent respectively. Moreover, in case of unfavorable health outcomes, the levels of CHC and FL turn out to be 43.3 percent and 41.6 percent respectively. Geographically, older workers are more concentrated in South, East and Central regions of India.

**Table-2:** Background characteristics of older workers (60 years and above), N=10,746 (34.16%)

| **Health outcomes: Type of work; Covariates** | **Percentage** |
| --- | --- |
| **Health Outcomes** |  |
| **Chronic Health Conditions** |  |
| No | 56.64 |
| Yes | 43.36 |
| **Functional Limitations** |  |
| No | 58.35 |
| Yes | 41.65 |
| **Poor Cognitive Functioning** |  |
| No | 70.87 |
| Yes | 29.13 |
| **Main predictor** |  |
| **Type of work** |  |
| Formal | 26.88 |
| Informal | 73.12 |
| **Socio-economic and demographic indicators** |  |
| **Gender** |  |
| Male | 67.59 |
| Female | 32.41 |
| **Age groups** |  |
| 60-65 | 54.01 |
| 65+ | 45.99 |
| **Caste groups** |  |
| General | 21.53 |
| Scheduled Tribe | 10.91 |
| Scheduled Caste | 21.17 |
| Other Backward Class | 46.39 |
| **Religion** |  |
| Hindu | 82.83 |
| Muslim | 11.10 |
| Others | 6.07 |
| **Education level** |  |
| Low | 78.17 |
| Middle | 16.30 |
| High | 5.52 |
| **Marital status** |  |
| Currently married | 74.84 |
| Others | 25.16 |
| **Place of residence** |  |
| Rural | 78.88 |
| Urban | 21.12 |
| **Wealth** |  |
| Low | 40.73 |
| Medium | 34.92 |
| High | 24.35 |
| **Household size** |  |
| 1 | 5.53 |
| 2 | 21.91 |
| 3 | 11.22 |
| 4+ | 61.34 |
| **Work Characteristics** |  |
| **Working hours** |  |
| Less than 24 hours | 27.49 |
| 24-48 hours | 42.19 |
| 48+ hours | 30.31 |
| **Average wage** | 5727.70 |
| **Life-style Behaviour** |  |
| **Drinking Alcohol** |  |
| No | 86.94 |
| Yes | 13.06 |
| **Smoking/Consuming Tobacco** |  |
| No | 49.13 |
| Yes | 50.87 |
| **Physical Activity** |  |
| **Vigorous** |  |
| Never | 42.28 |
| Rare | 21.38 |
| Everyday | 36.34 |
| **Moderate** |  |
| Never | 29.32 |
| Rare | 15.09 |
| Everyday | 55.59 |
| **Yoga/Pranayam** |  |
| Never | 87.60 |
| Rare | 4.55 |
| Everyday | 7.85 |
| **Regions** |  |
| North | 9.14 |
| Central | 20.04 |
| East | 23.44 |
| Northeast | 2.76 |
| West | 19.09 |
| South | 25.35 |
| Union Territories | 0.17 |

### Bivariate analysis results

When health outcomes were examined by type of work, as shown in Fig 2, formal older workers suffer from high burden of CHC as compared to informal counterparts (47.1% vs. 41.9%, p < 0.0001). On the other hand, the risk of FL and PCF is more prevalent among informal workers (FL: 42.5% vs. 39.3%, p < 0.05; PCF: 32.5% vs. 20.0%, p < 0.0001).

**Fig 2:**
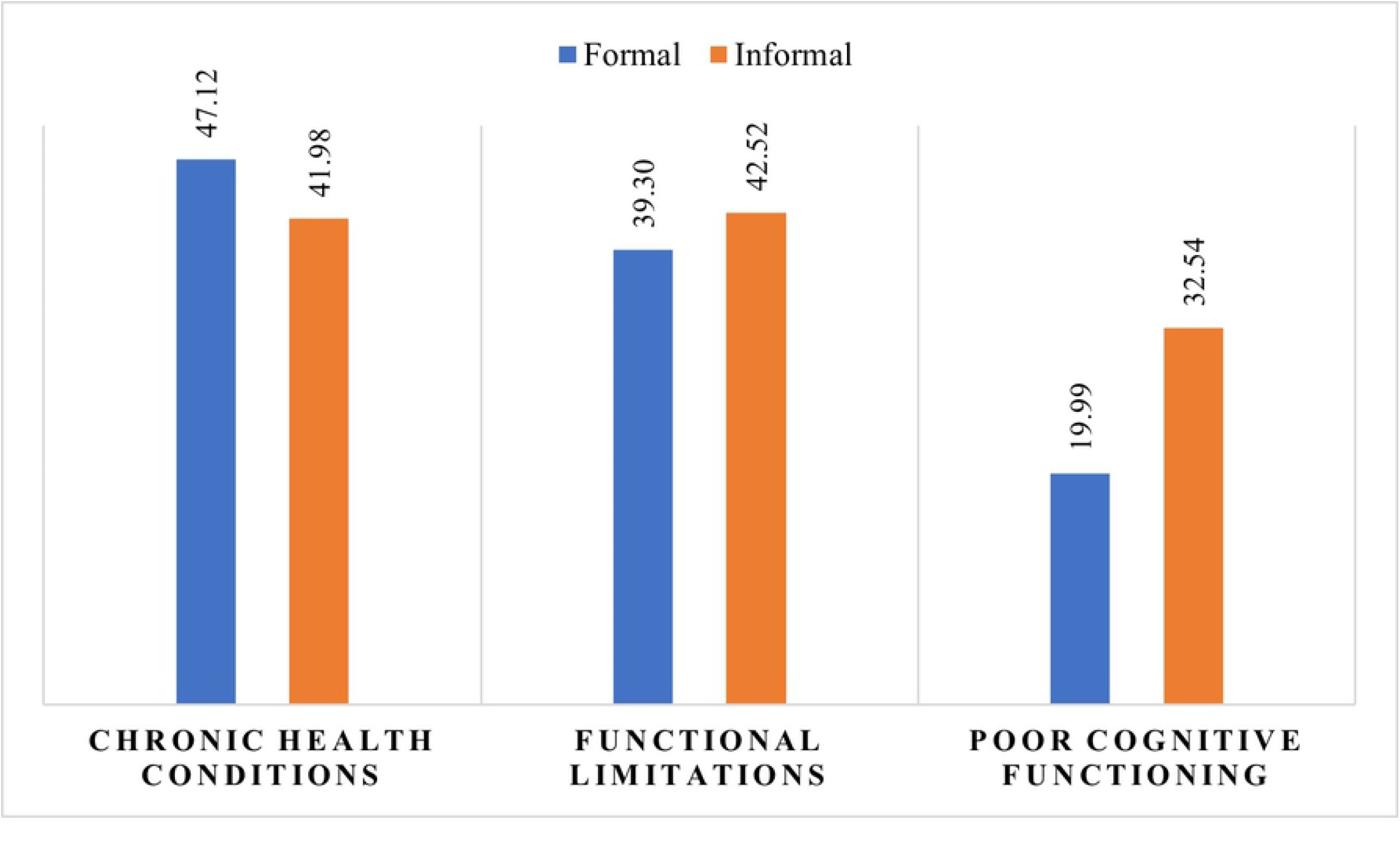
Level of health outcomes by type of work.

Table 3 presents the estimated rates of sampled older workforce of LASI data by the type of work they engaged in. From the table, it is observed that majority of female workforce are engaged in informal activities, conversely among male population major share lies in formal activities. In context of caste groups, the rate for informal activities is high among SC and ST, while general and OBC holds notable portion in formal activities. The percentage for informal workers is substantial among Muslim and other communities compared to Hindu community. Education and wealth play a significant role in defining nature of work. Low level of financial wellbeing and education reflects high share of work engagement in informal activities. As expected, the level of informal older workers is quite considerable in rural areas, whereas urban areas have a significant share of formal workers. Those who are living alone tend to work in informal activities that those living with family members. Besides, concentration of informal workers is more in North-East followed by South and Western regions of India. Further, marital status and working hours reflects a meagre difference in determining type of work.

**Table-3:**
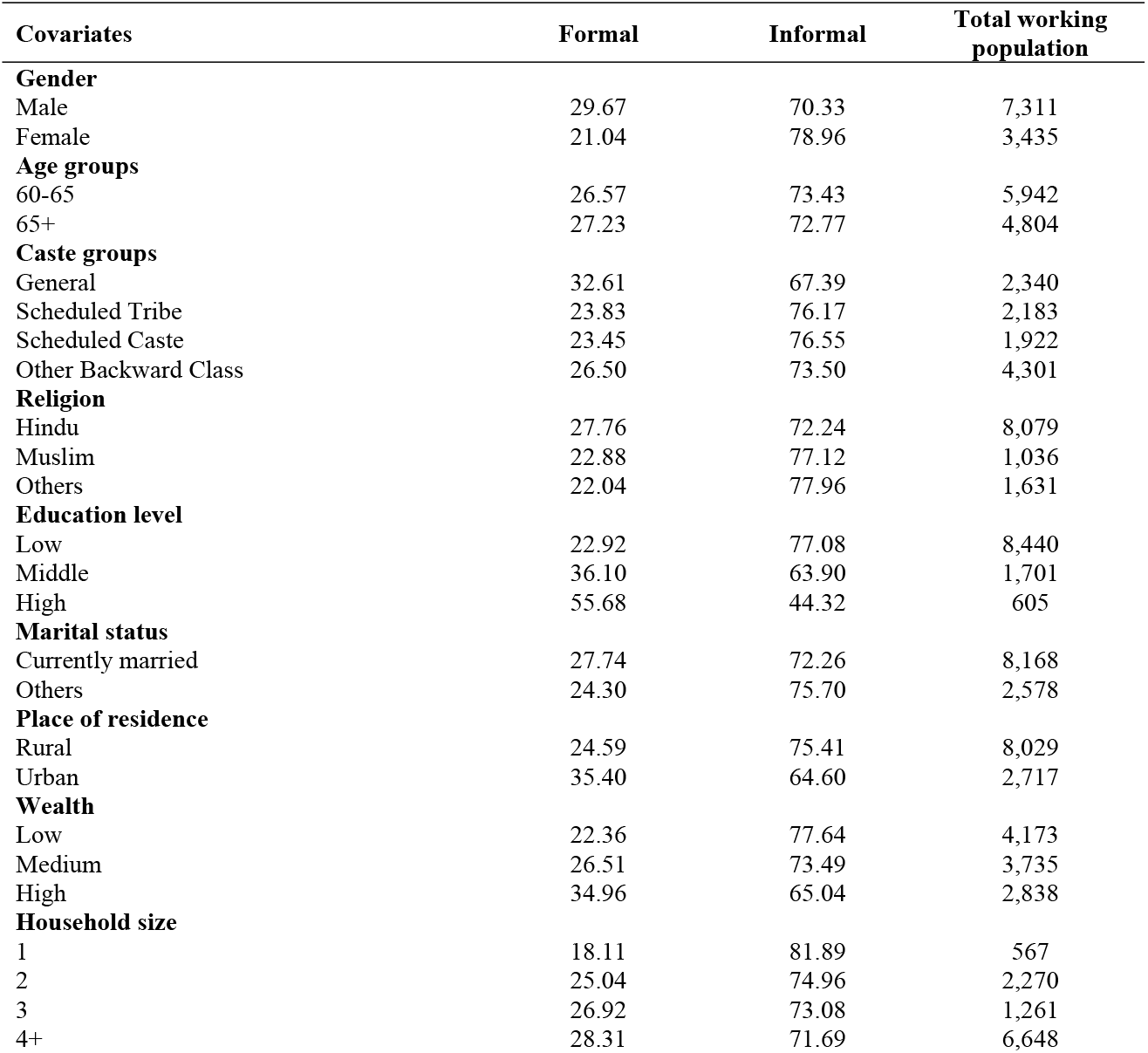
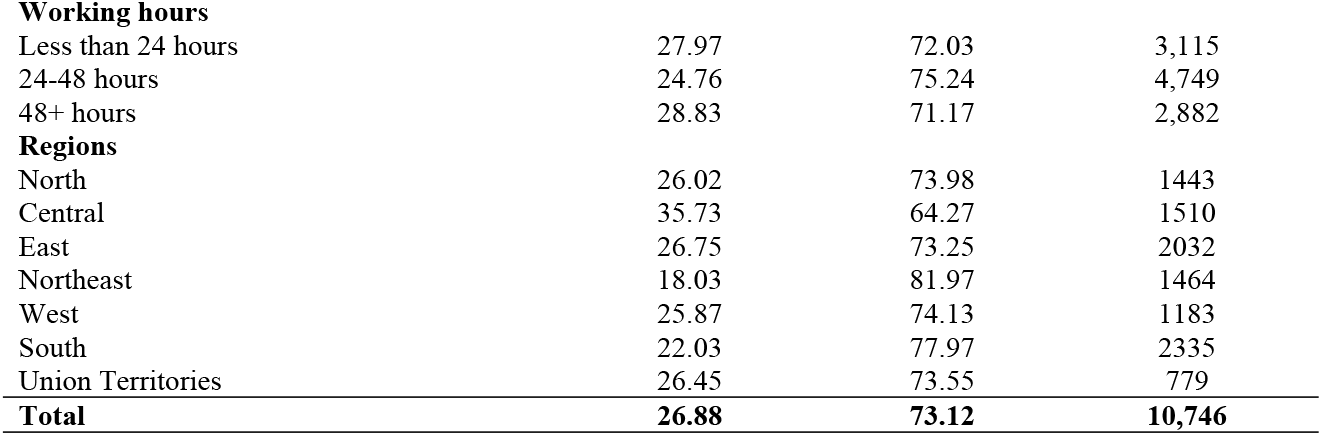
Type of work by socio-economic and demographic characteristics.

| **Covariates** | **Formal** | **Informal** | **Total working population** |
| --- | --- | --- | --- |
| **Gender** |  |  |  |
| Male | 29.67 | 70.33 | 7,311 |
| Female | 21.04 | 78.96 | 3,435 |
| **Age groups** |  |  |  |
| 60-65 | 26.57 | 73.43 | 5,942 |
| 65+ | 27.23 | 72.77 | 4,804 |
| **Caste groups** |  |  |  |
| General | 32.61 | 67.39 | 2,340 |
| Scheduled Tribe | 23.83 | 76.17 | 2,183 |
| Scheduled Caste | 23.45 | 76.55 | 1,922 |
| Other Backward Class | 26.50 | 73.50 | 4,301 |
| **Religion** |  |  |  |
| Hindu | 27.76 | 72.24 | 8,079 |
| Muslim | 22.88 | 77.12 | 1,036 |
| Others | 22.04 | 77.96 | 1,631 |
| **Education level** |  |  |  |
| Low | 22.92 | 77.08 | 8,440 |
| Middle | 36.10 | 63.90 | 1,701 |
| High | 55.68 | 44.32 | 605 |
| **Marital status** |  |  |  |
| Currently married | 27.74 | 72.26 | 8,168 |
| Others | 24.30 | 75.70 | 2,578 |
| **Place of residence** |  |  |  |
| Rural | 24.59 | 75.41 | 8,029 |
| Urban | 35.40 | 64.60 | 2,717 |
| **Wealth** |  |  |  |
| Low | 22.36 | 77.64 | 4,173 |
| Medium | 26.51 | 73.49 | 3,735 |
| High | 34.96 | 65.04 | 2,838 |
| **Household size** |  |  |  |
| 1 | 18.11 | 81.89 | 567 |
| 2 | 25.04 | 74.96 | 2,270 |
| 3 | 26.92 | 73.08 | 1,261 |
| 4+ | 28.31 | 71.69 | 6,648 |
| **Working hours** |  |  |  |
| Less than 24 hours | 27.97 | 72.03 | 3,115 |
| 24-48 hours | 24.76 | 75.24 | 4,749 |
| 48+ hours | 28.83 | 71.17 | 2,882 |
| **Regions** |  |  |  |
| North | 26.02 | 73.98 | 1443 |
| Central | 35.73 | 64.27 | 1510 |
| East | 26.75 | 73.25 | 2032 |
| Northeast | 18.03 | 81.97 | 1464 |
| West | 25.87 | 74.13 | 1183 |
| South | 22.03 | 77.97 | 2335 |
| Union Territories | 26.45 | 73.55 | 779 |
| **Total** | **26.88** | **73.12** | **10,746** |

### Chronic Health Conditions

Table 4 depicts the risk of CHC among older working population in India (crude and adjusted). Informal workers tend to have less odds of CHC compared to formal counterparts. Indeed, their odds are 0.82 folds less (OR=0.816, 95% CI: 0.748-0.890, p<0.0001). The odds ratios change slightly and remain significant after controlling socio-economic and demographic variables (in Model-2), and other covariates (in Model-3).

**Table-4:**
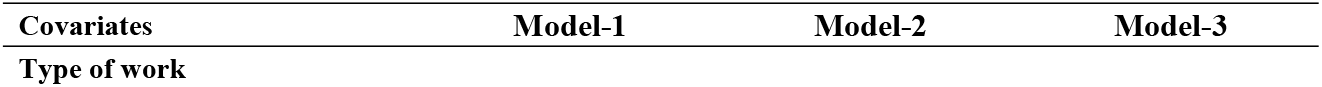
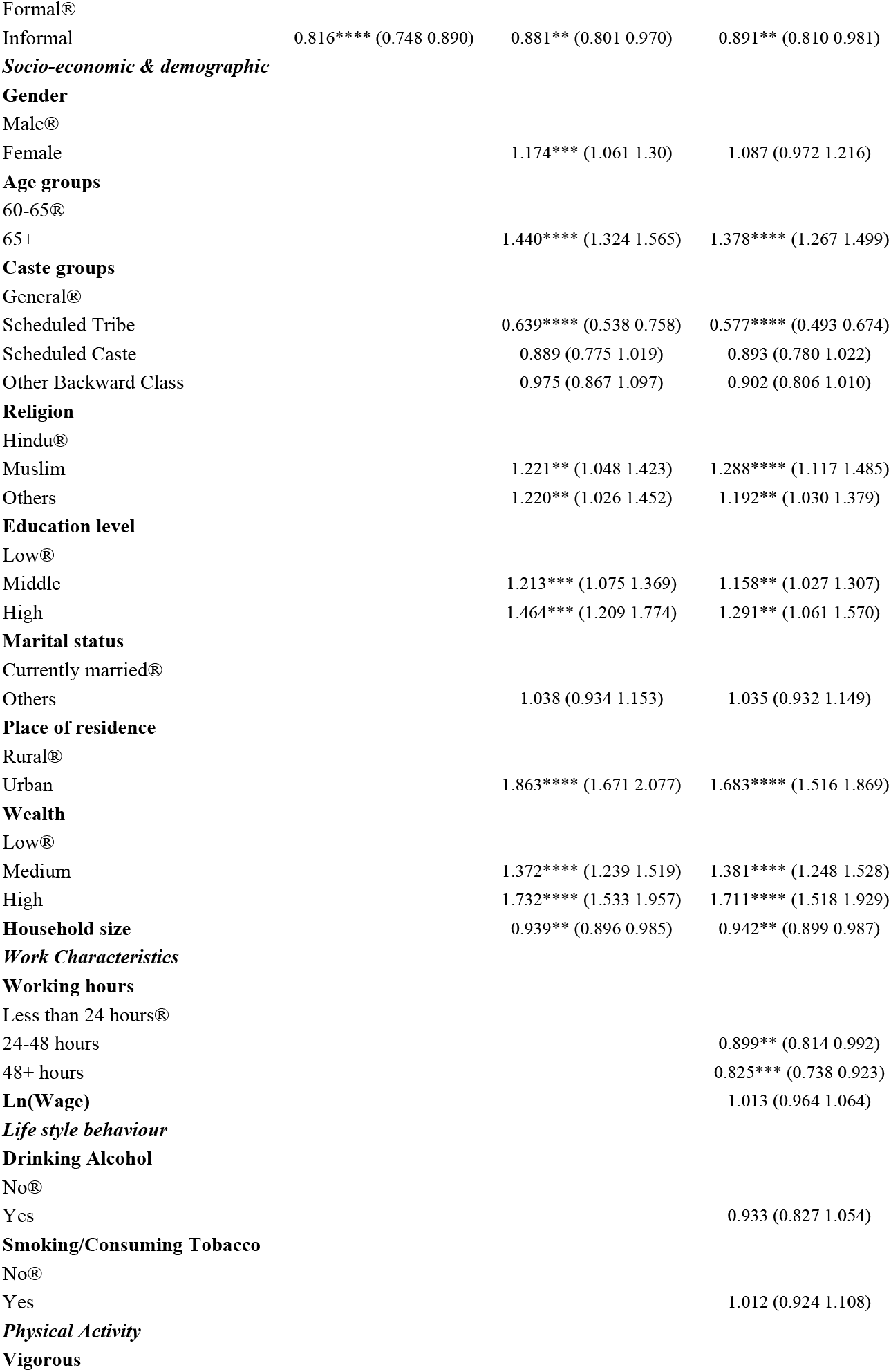
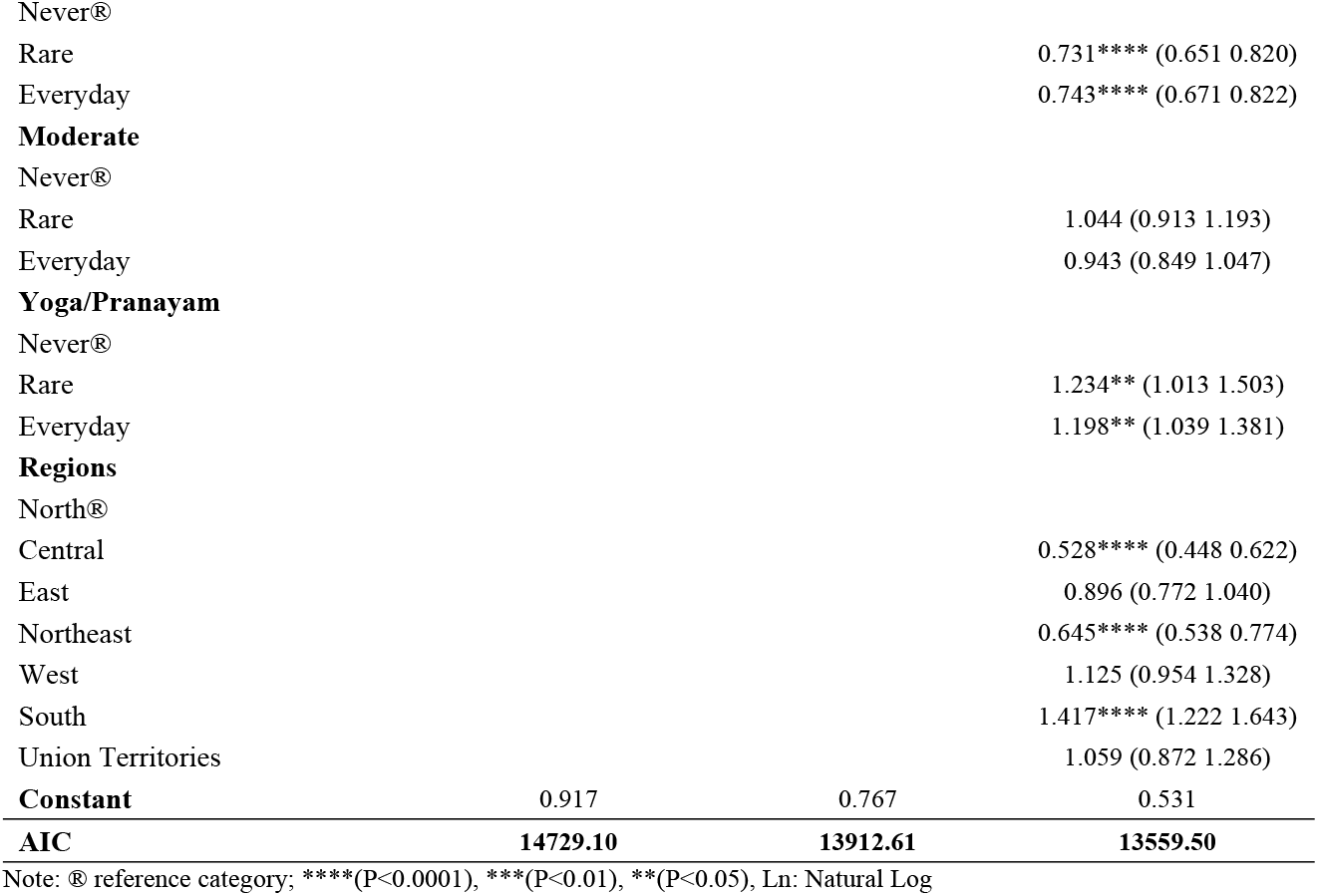
Result of multiple logistic regression for CHC.

Additionally, among the many covariates, the model indicates that 65+ age-group is associated with increased risk of CHC. The likelihood of CHC is significantly low among older workers from ST and OBC groups. On the other hand, this risk expands for Muslim and other religious communities. The odds of CHC significantly inflates with the increase in education and wealth. Similarly, urban workers are having 1.683 times (OR=1.683, 95%CI: 1.516-1.869, p<0.0001) more odds of CHC than rural older workers. The long working hours is significantly associated with low risk of CHC. Moreover, the physical activities do have significant influence over chronic conditions but with varying directions. Vigorous activities tend to have low risk of CHC, while those who perform Yoga/Pranayam on daily basis have 1.198 times (p<0.05) more odds of chronic conditions. Across India, the likelihood of CHC considerably high in South region, while it is low in Central and North-Eastern regions as compared to North.

### Functional Limitations

Table 5 exhibits the relationships between FL, CHC and type of work, and shows that both CHC and type of work are significant risk factors for FL. From Model-1, the odds of FL are 1.129 times more among informal workers in contrast to formal counterparts (p<0.01) after controlling for CHC, while the odds are 1.465 times more among those suffering CHC (p<0.0001) after controlling for the type of work. Further, interacting CHC with type of work (Model-2 & Model-3) reveals that both type of work and CHC maintain their significant level. Indeed, informal workers without CHC have 0.8 folds less odds of FL in contrast to formal workers without CHC, adjusted for all the covariates (OR=0.852, 95% CI: 0.764-0.951, p<0.001). For older workers with CHC, the odds of FL due to the type of work are quite similar in magnitude (OR=0.985, 95% CI: 0.799-1.216, p<0.001). However, Model-2 and Model-3 interaction results show that the type of work does significantly influence the effect of CHC on FL. Indeed, as shown in Model-3, for formal workers, the odds of FL are 1.480 (p<0.0001) times higher among those with CHC. For informal workers with CHC, the odds ratio expands by 1.712 times (OR: 1.157*1.480, 95% CI: 1.414-2.073, p<0.001) compared to informal without CHC.

**Table-5:**
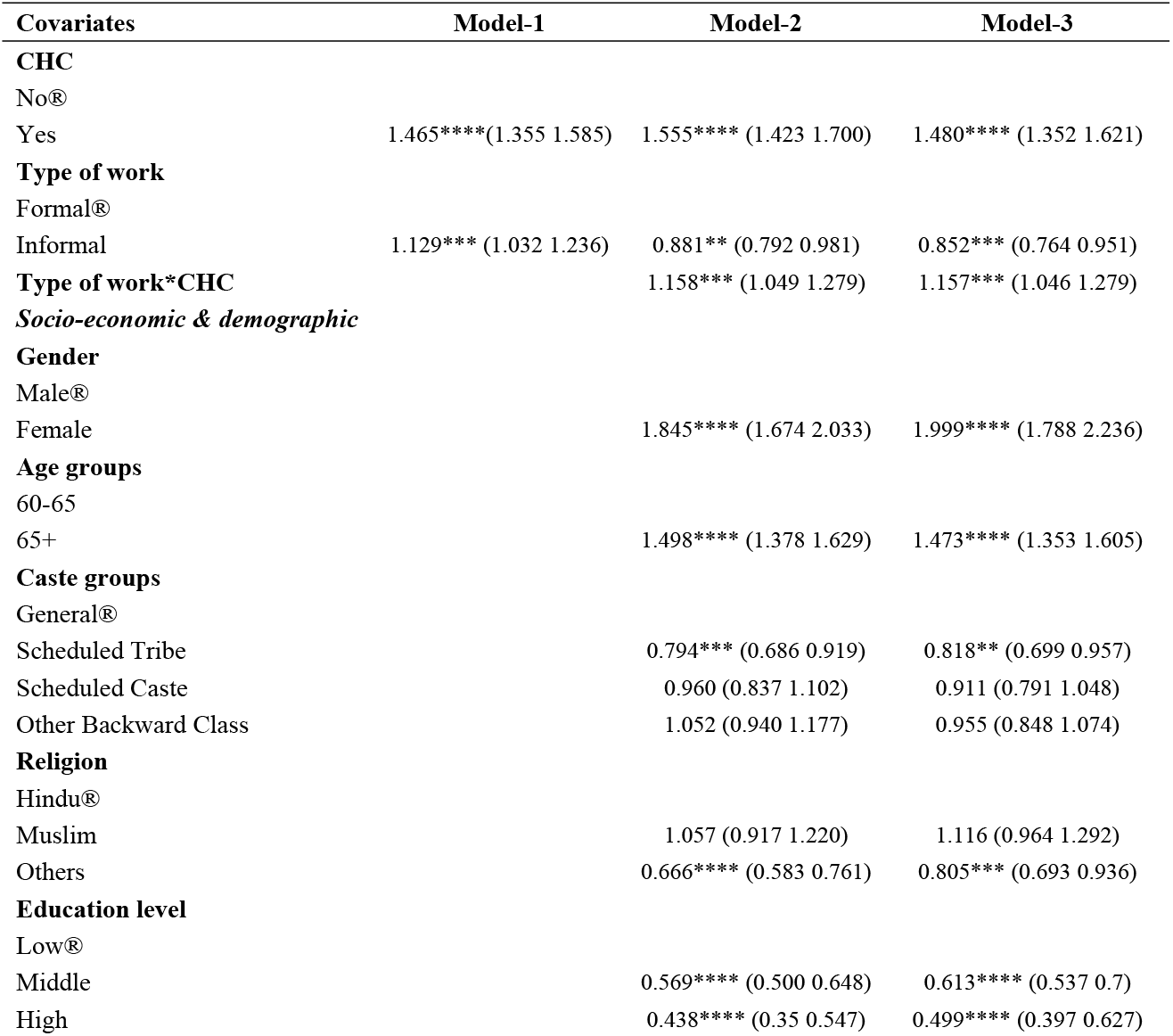
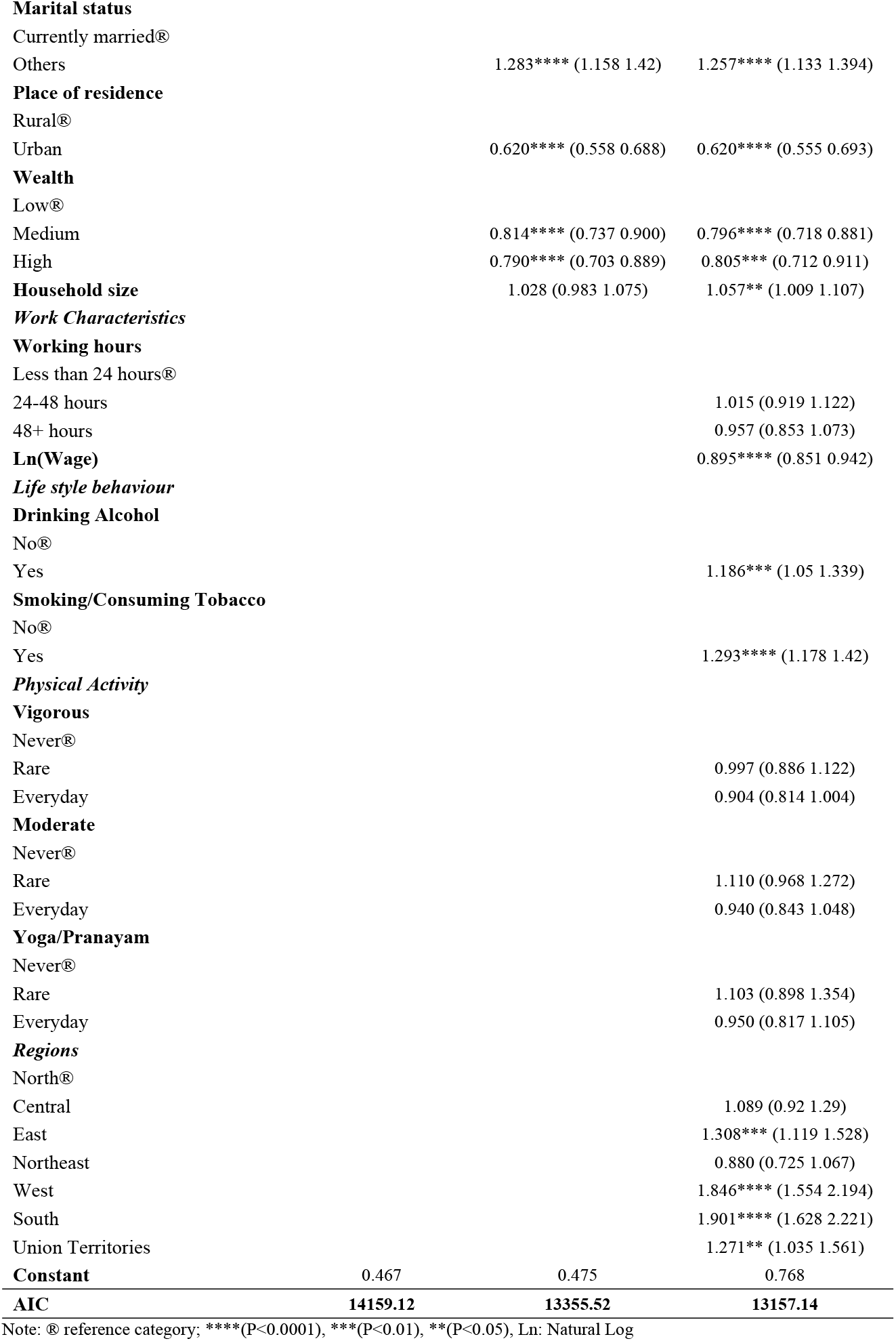
Result of multiple logistic regression for FL.

Apart from these key results, among all covariates, it appeared that females are more prone to FL, while educated, wealthy and ST population are less likely to suffer from the same. Further, high odds of FL are common among those who are engaged in unhealthy lifestyle behaviours such as drinking alcohol and consuming tobacco or smoking. Besides, the odds of FL are 1.257 (p<0.0001) times greater among those who are not currently married. Finally, across India, the odds of FL are relatively considerable in Southern and Western regions compared to North.

### Poor Cognitive Functioning

From Table 6, it is observed that the type of work, FL and CHC are significantly associated with PCF after controlling for each other (Model-1). Indeed, the odds of PCF (adjusted for CHC and type of work) are 2.14 folds among workers with FL (OR=2.140, 95%CI: 1.961-2.336, p < 0.0001; compared to non-FL). Moreover, the odds of PCF (adjusted for CHC and FL) are 1.7 folds among informal workers (OR=1.700, 95%CI: 1.528-1.892, p < 0.0001; compared to formal). Finally, the odds of PCF (adjusted for FL and type of work) are 0.77 folds among workers with CHC (OR=0.766, 95%CI: 0.701-0.836, p < 0.0001; compared to non-CHC).

**Table-6:**
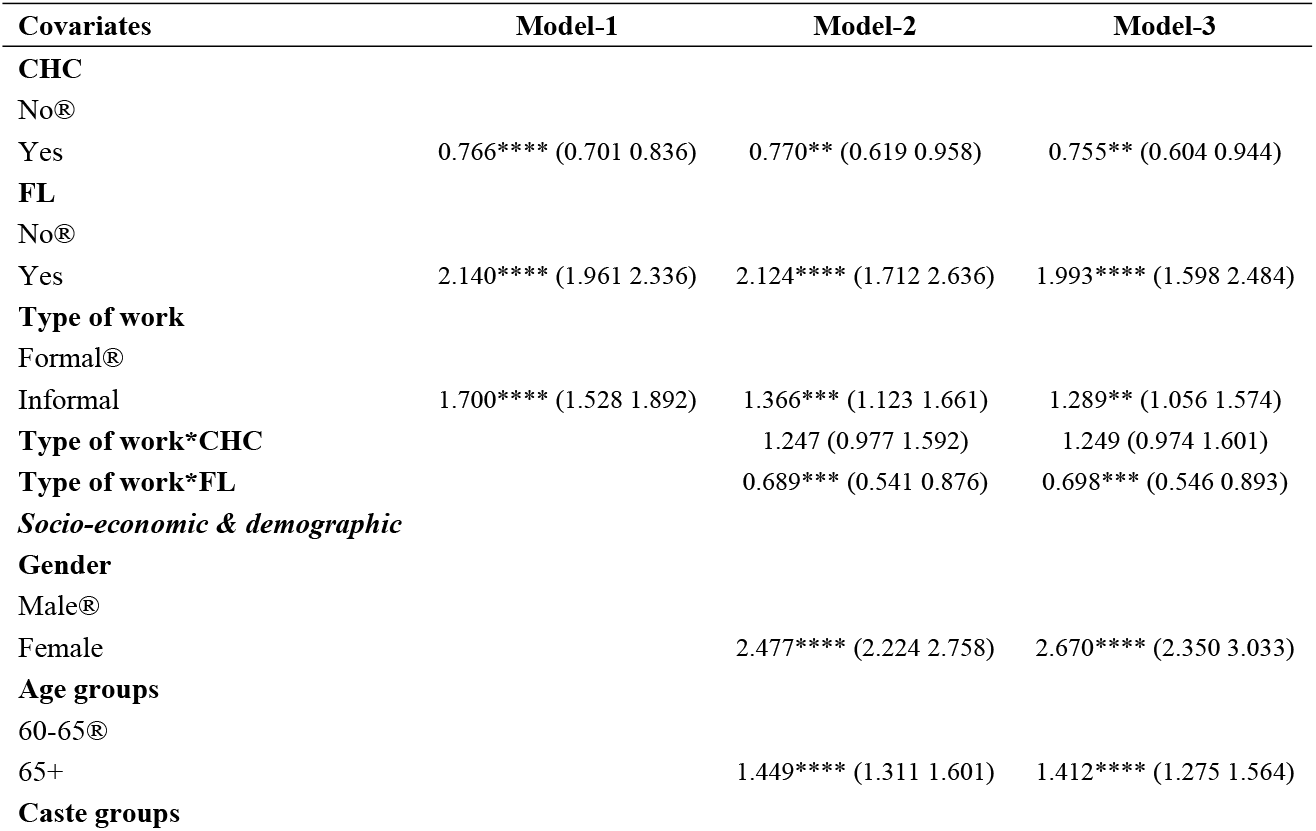
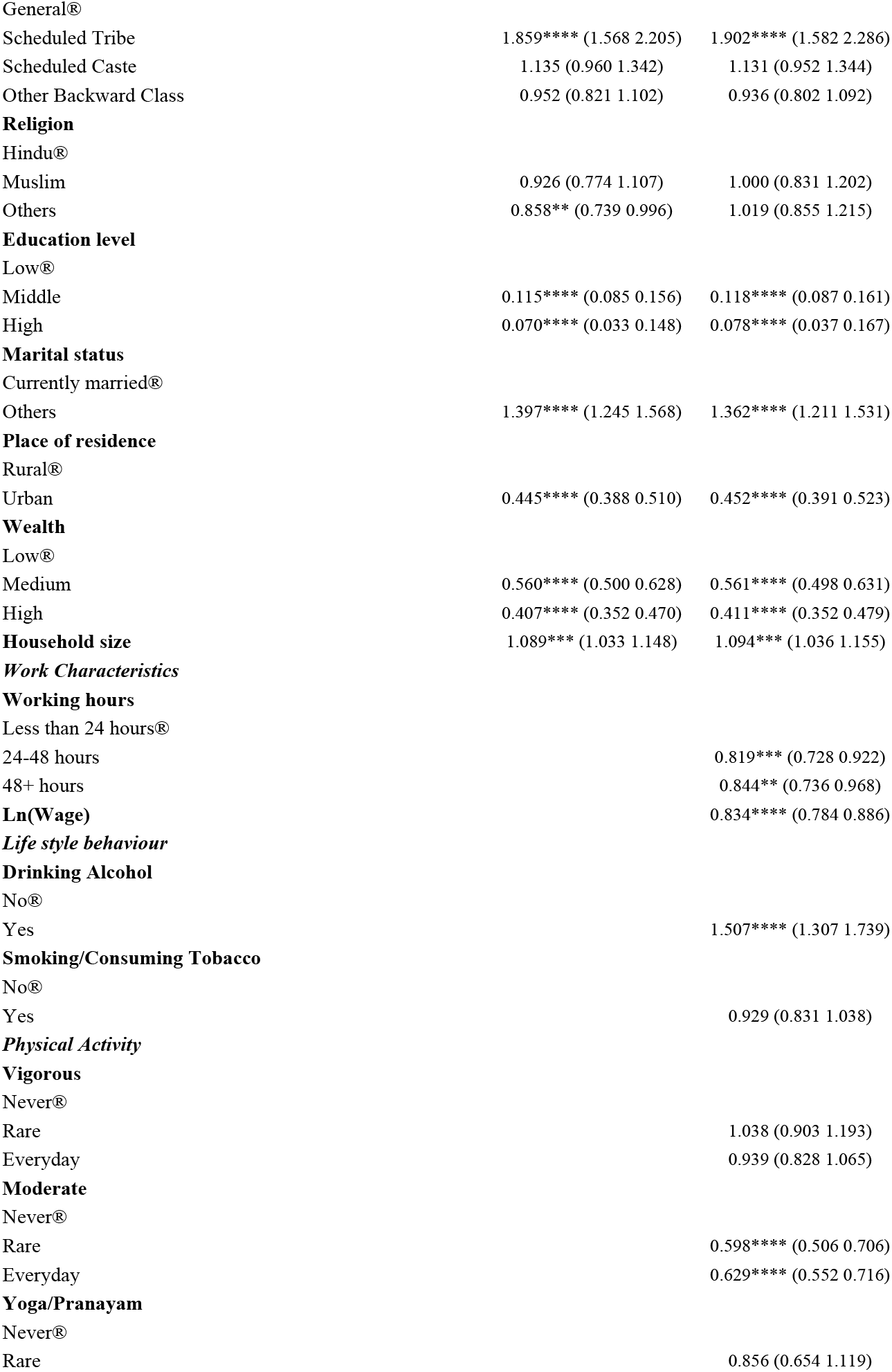
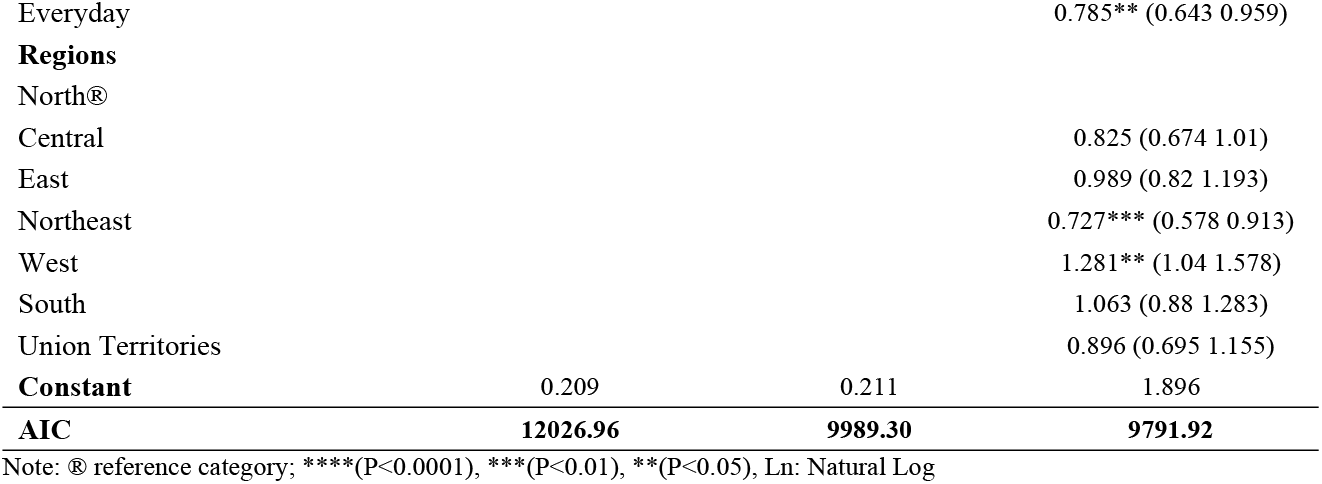
Result of multiple logistic regression for PCF.

In Model-3, adjusting for all covariates, demonstrates that formal workers with CHC have 0.76 times less odds of PCF in contrast to formal workers without any CHC (OR=0.755, 95%CI: 0.604-0.944, p<0.05). Likewise, formal workers with FL have 1.993 times odds of PCF compared to formal workers without any FL (OR=1.993, 95%CI: 1.598-2.484, p<0.0001). Further, the odds of PCF are 1.289 times for informal workers without any poor physical health condition (no CHC and no FL) as compared formal workers without any poor physical health condition (OR=1.289, 95%CI: 1.056-1.574, p<0.05). Additionally, the type of work moderates significantly the effect of FL on PCF, but not significantly the effect of CHC (see interaction effects as differential factors in Model-2 and Model-3). Indeed, as shown in Model-3, informal workers with any FL are 1.391 times (OR=1.993*0.698, 95%CI: 0.872-2.218, p<0.001) more prone to PCF than informal workers without FL. Likewise, informal workers with any CHC are 0.943 times (OR=0.755*1.249, 95%CI: 0.588-1.511, p<0.05) less prone to PCF than informal workers without CHC. Although the differential factor is statistically non-significant, the odds ratio (informal workers with CHC compared to no CHC) are statistically significant.

On the other hand, the type of work on PCF is modified, to a certain extent, by CHC and FL. Indeed, informal workers with CHC have 1.609 times (OR=1.249*1.289, 95%CI: 1.028-2.519, p<0.05) more odds of PCF compared with formal worker with CHC. Conversely, the odds of PCF are 0.900 times (OR=0.698*1.289, 95%CI: 0.576-1.406, p<0.05) less among informal workers with any FL compared to their formal counterparts with FL.

Among all covariates, a noteworthy gap can be seen among female workers in the odds of PCF when compared to male workers (Model-3). Likewise, the high odds of PCF are significant among ST groups, 65+ cohort and currently not married population as well as those who consume alcohol. The odds of PCF increase as well with the increase in household size. On the other hand, the risk decreases with rise in wealth, educational level, working hours, wage, and movement from rural to urban areas as shown by the estimated odds ratios. Nevertheless, physical activities play important role in improving cognitive function as evident from the odds of moderate exercise and Yoga/Pranayam. Geographically, the risk of PCF is high as indicated by the estimated odds ratio (OR=1.281, p<0.05) in West region and low (OR=0.727, p<0.001) in North-Eastern region in comparison to North.

## Discussion

In the backdrop of increasing older population in India and the paucity of pension/financial benefits schemes, it is quite likely that this population is still working after the retirement age. The present study supports this argument and observed that around one-third of the population aged 60 years and above are in labour force. However, the majority of this population are engaged in informal activities contributing nearly 73 percent of total older population. Besides, the share of labour force participation is higher among males than that of females. On the other hand, the participation rate of female population in informal activities is greater compared to male counterparts. Certainly, females are generally expected to spend most of the time in household activities and taking care of their family members. Consequently, they choose low paid elementary activities which needs less work time (2, 20, 51, 52). Likewise, the engagement in informal work is highest among Muslim community, Scheduled Caste, and Scheduled Tribe groups. It is well documented that these sections of the society in India are marginalized due to their poor socio-economic conditions. Reddy (20) explains that socio-economically disadvantage populations tend to have high labour-force participation rate in later ages. Further, pronounced level of participation in can be observed among older population with low education and wealth. This large workforce is mostly engaged in agricultural activities, casual labour or unskilled occupations which require less education (2, 20). Generally, agricultural activities in India are concentrated in rural areas and involve subsistence farming, resulting in high proportion of workforce participation in later life. Consequently, agrarian households tend to have larger family size which is assumed to provide better manpower to support farming activities. This can be clearly seen through current findings showing higher work participation for larger family size households. However, it is also important to note that the level of engagement in informal activities is considerable for those households with small family sizes. Tafuro (66) elucidated that intergenerational support is strongly related with son’s preference in countries like, India, China, Vietnam, and South Korea, and it is quite common that older people without sons are more likely to be economically active in these regions.

Previous studies demonstrated that work engagement has a pronounced effect on physical and mental health. Consistent with these articles, the present research reaffirms that work engagement does play a significant role in determining unfavourable health outcomes. Nevertheless, current findings also provide the evidence of varied unfavourable health outcomes by type of economic activity which has not been documented in former research. In fact, informal older workers are less likely to suffer from CHC and FL, while more likely to have PCF as compared to formal older workers. The emergence of high CHC rates is most likely to be observed among educated and wealthy groups (24, 26, 67) which is closely attributed to socio-economic profiles of formal workers. These relationships have retained their statistical significance level even after controlling for all the covariates in each model. Nonetheless, noticeable change in nature of relationship is observed between type of work and FL. In model-1, the risk of FL is substantially high among informal workers but, after controlling for the covariates in model-3, the risk of FL changed drastically reflecting favourable effect of informal activities on FL for older people with no chronic health conditions. The low risk of FL among informal workers confirms healthy work effect concept suggested by former literatures (48, 52, 57, 68) because informal activities involve physically demanding works and only those can be engaged in later life who are not suffering from severe physical functioning. However, the relationship between type of work and FL is influenced by CHC because informal workers without any chronic conditions have 15 percent less risk of FL as compared to formal workers, whereas this percentage gap decreases to 2 percent for those who suffers from any CHC. Empirical evidence explains that the burden of CHC amplifies functional limitations and makes it quite cumbersome to remain active in later ages (16, 69).

Continuous work engagement in later life may have leading implication on cognitive functioning (11, 48, 50, 51). Extending from this relationship, the present research also illustrates varied influence of type of work on PCF. For instance, informal workers without any physical health condition (e.g., CHC and FL) are more likely to suffer from PCF as compared to formal workers without any health condition. Intriguingly, this risk elevates by 1.609 times for informal workers with CHC in contrast to formal workers with CHC which shows that CHC may influence the relationship between type of work and PCF. This can be explained by the fact that informal older workers have inadequate financial resources to seek treatment for CHC, and due to absence of social and financial protection schemes, they suffer from serious loss of mental or physical ability (18-20, 42). Nonetheless, the PCF turns out to be higher among those formal workers who have any FL when equated to informal workers with FL. Present results provide an interesting insight that informal with CHC have negative ramification on cognitive functioning, while formal with FL have adverse effect on cognitive functioning. Formal older workers generally belong to those sectors where brainstorming is required, therefore having FL will have a larger implication on PCF among this group because it will reduce their mental capacity to work. Besides, it has been established from certain studies that FL is a key role player in shaping cognitive ability (49, 51). Altogether, summarising the relationship between type of work and health outcomes, it can be suggested that FL is more damaging for formal workers because it deteriorates their cognitive functioning, whereas for informal workers CHC is a crucial factor in hampering physical functioning.

This research also sheds light on those determinants which has considerable impact on unfavourable health outcomes. Those are namely socio-economic and demographic attributes, work characteristics, lifestyle behaviour, and regions of India. Among them, prolonged work engagement has detrimental effect on physical and cognitive functioning for those who are female older workers if likened to male counterparts. Commonly mainstream female workers are employed in informal activities, and they alone work as domestic workers giving rise to double burden of activities (51, 52). Additionally, it is known that women live longer than men and continue to work in their later life with one or more disabilities (11, 52, 70). In Indian society, ST and Muslim community are also the part of vulnerable and marginalized sections who tend to work longer in later ages. The risk of PCF is substantial among ST groups, while the burden of CHC is more prominent among Muslim community. This is may be because ST group is mostly engage in manual labour or scavenging (71), and agricultural labour activities (72) which is associated with decline in cognitive ability (51). On the other hand, Muslim community in India are well known for their engagement in informal artisanal work such as weaving, carpentry, black-smithing, and Zari work which is related ergonomic condition leading to CHC (73-76).

Education and wealth are considerable predictors of unfavourable health outcomes. Increase in education and wealth leads to reduction in FL and PCF, though escalating the chances of CHC. As evident from studies that better education and wealth improves the physical functioning and work ability (44, 45). In case of work characteristics, increase in working hours and wages is associated with improvement in PCF, but this result is contradictory to the studies conducted in Japan which states that part time or low working hours improves the cognitive functioning of older workers (10, 11). It is because Japan is a developed country where provision for social and financial security is strong, therefore older workers only engage to maintain their social network and get the sense of pride of being economically independent. The life-style behaviours are essential components which needs to be taken into account to justify the relationship between type of work and health. Drinking alcohol and consuming tobacco/smoking can lead to serious health implication, as evident from the result that it degrades physical functioning. Subsequently, vigorous physical exercise diminishes the risk of CHC and FL, however Yoga/Pranayam ameliorates cognitive functioning. Moreover, adverse impacts of CHC and FL can be minimized by staying active and maintaining a healthy lifestyle (77-79). Heterogeneity in context of health outcomes across regions can be clearly observed in India. Notably, chances of CHC upsurges in central and north-eastern region, but FL is relatively intensifying in southern and western region. Geographically, there is a stark difference in terms of PCF, it rises in western region, then dwindles in north-eastern region. Despite of physical and mental health limitations, older population in India are still working after the retirement age especially in informal sector.

Further, this study has certain limitations such as use of cross-sectional data at a single point of time. Thus, result of the study could only provide the evidence of statistical association between type of work and health outcomes not the cause-effect relationship. To get the better picture of type of work and health, longitudinal data would be a better option. However, there is no longitudinal data which focuses on older people in India.

## Conclusion

Older population in India constitute an undeniable share of labour-force after the retirement age of 60 years. This extent is the outcome of financial constraints caused by paucity in social and health insurance schemes. Moreover, working longer impacts physical and mental health of the older people which varies by formal or informal sector of employment. Further, improving health conditions of this vulnerable population should be an utmost priority for policy makers to encourage active and healthy ageing.

Therefore, present study underscores the relevance of policies focusing on providing health and healthcare benefits by respective economic activity and socio-economic position. Adequate working conditions should be considered during policy formulation, which can offer a level of job satisfaction and contribute to better well-being. Moreover, the preference of employment type should be given precedence according to the age-groups. It would be better that older adults aged 70+ should participate in part-time jobs as they may be unsuitable to handle high strain and physically demanding work. Economic security policy should be recommended to those who are lacking physical capacity. This can help them to sustain their livelihood without any financial constraints. The issues of older workers in India should be taken seriously otherwise it will lead to a huge chunk of vulnerable groups with inadequate social and financial support.

## Data Availability

Data are available from International Institute for Population Sciences (IIPS) - contact via

https://iipsindia.ac.in/content/LASI-data

## Acknowledgements

I humbly thanks International Institute for Population Sciences (IIPS) for giving me access to Longitudinal Ageing Study of India dataset for this research.

## Contributors

PC contributed to conceptualization, methodology, data analysis and writing the original draft. TN and IM contributed to conceptualization, methodology, supervision, and writing (review and editing). AS contributed to validation and writing (review and editing).

## Ethical Approval

This research paper is based on secondary dataset, which is available in public domain, however, before using the dataset approval has been taken from IIPS.

## Data Availability Statement

Data is available in public domain. The link is : https://iipsindia.ac.in/content/LASI-data

## References

1. Christensen K, Doblhammer G, Rau R, Vaupel JW. Ageing populations: the challenges ahead. The Lancet. 2009;374(9696):1196–208.

2. Adhikari R, Soonthorndhada K, Haseen F. Labor force participation in later life: evidence from a cross-sectional study in Thailand. BMC Geriatr. 2011;11:15.

3. World Health Organization. Aging and health 2018 [updated 5 February 2018. Available from: http://www.who.int/news-room/fact-sheets/detail/ageing-and-health.

4. McAllister A, Bentley L, Bronnum-Hansen H, Jensen NK, Nylen L, Andersen I, et al. Inequalities in employment rates among older men and women in Canada, Denmark, Sweden and the UK. BMC Public Health. 2019;19(1):319.

5. Larsen M, Pedersen PJ. Labour force activity after 65: what explain recent trends in Denmark, Germany and Sweden? J Labour Mark Res. 2017;50(1):15–27.

6. Nilsson K, Ostergren PO, Kadefors R, Albin M. Has the participation of older employees in the workforce increased? Study of the total Swedish population regarding exit from working life. Scand J Public Health. 2016;44(5):506–16.

7. Kalwij A, Kapteyn A, De Vos K. Work capacity at older ages in the Netherlands. National Bureau of Economic Research; 2016. Report No.: 0898-2937.

8. Garcia MTM, Fontainha E, Passos J. Hiring older workers: The case of Portugal. The Journal of the Economics of Ageing. 2017;9:71–7.

9. Temple JB, Rice JM, McDonald PF. Mature age labour force participation and the life cycle deficit in Australia: 1981–82 to 2009–10. The Journal of the Economics of Ageing. 2017;10:21–33.

10. Minami U, Nishi M, Fukaya T, Hasebe M, Nonaka K, Koike T, et al. Effects of the Change in Working Status on the Health of Older People in Japan. PLoS One. 2015;10(12):e0144069.

11. Tomioka K, Kurumatani N, Hosoi H. Beneficial effects of working later in life on the health of community-dwelling older adults. Geriatr Gerontol Int. 2018;18(2):308–14.

12. Lee J, Kim MH. The effect of employment transitions on physical health among the elderly in South Korea: A longitudinal analysis of the Korean Retirement and Income Study. Soc Sci Med. 2017;181:122–30.

13. Jousten A, Lefebvre M. Work capacity and longer working lives in Belgium. National Bureau of Economic Research; 2016. Report No.: 0898-2937.

14. UNITED NATIONS. World Population Ageing 2017-Highlights. Department of Economic and Social Affairs. 2017.

15. ILO. World social protection report 2017–19: Universal social protection to achieve the Sustainable Development Goals. 2017.

16. Dantas RG, Perracini MR, Guerra RO, Ferriolli E, Dias RC, Padula RS. What are the sociodemographic and health determinants for older adults continue to participate in work? Arch Gerontol Geriatr. 2017;71:136–41.

17. Liu H, Lou WQ. Patterns of productive activity engagement among older adults in urban China. Eur J Ageing. 2016;13(4):361–72.

18. Dhar A. Workforce participation among the elderly in India: Struggling for economic security. The Indian Journal of Labour Economics. 2014;57(3):221–45.

19. Rajan SI. Demographic ageing and employment in India. Bangkok: International Labour Organization, Regional Office for Asia and the Pacific(ILO Asia-Pacific Working paper series). 2010.

20. Reddy AB. Labour force participation of elderly in India: patterns and determinants. International Journal of Social Economics. 2016;43(5):502–16.

21. United Nations. World Population Ageing 2015. D. o. E., & Social Affairs, P. D. United Nations New York, NY. 2015.

22. United Nations. World Population Prospects 2019, Volume II: Demographic Profiles 2019 [Available from: https://population.un.org/wpp/Graphs/DemographicProfiles/Line/356.

23. Bose M, Banerjee S. Equity in distribution of public subsidy for noncommunicable diseases among the elderly in India: an application of benefit incidence analysis. BMC Public Health. 2019;19(1):1735.

24. Ingle GK, Nath A. Geriatric health in India: concerns and solutions. Indian J Community Med. 2008;33(4):214–8.

25. Dey S, Nambiar D, Lakshmi J, Sheikh K, Reddy KS. Health of the elderly in India: challenges of access and affordability. Aging in Asia: Findings from new and emerging data initiatives: National Academies Press (US); 2012.

26. Mini GK, Thankappan KR. Pattern, correlates and implications of non-communicable disease multimorbidity among older adults in selected Indian states: a cross-sectional study. BMJ Open. 2017;7(3):e013529.

27. Chowdhury P, Garg MK, Ladusingh L. Morbidity Pattern of Elderly in India. Issues on Health and Healthcare in India: Springer; 2018. p. 445–58.

28. Talukdar B. Prevalence of multimorbidity (chronic NCDS) and associated determinants among elderly in India. Demographic India. 2017:69–76.

29. Agrawal G, Keshri K. Morbidity patterns and health care seeking behavior among older widows in India. PLoS One. 2014;9(4):e94295.

30. Shil A, Puri P, Prakash R. A geospatial analysis of noncommunicable disease (NCD) burden in Indian agro-climatic and political regions. Journal of Public Health. 2018;26(4):391–8.

31. Kowal P, Kahn K, Ng N, Naidoo N, Abdullah S, Bawah A, et al. Ageing and adult health status in eight lower-income countries: the INDEPTH WHO-SAGE collaboration. Global Health Action. 2010;3(1):5302.

32. World Health Organization. Health systems financing: The path to iniversal coverage. The World Health Report. 2010:1–128.

33. Pradhan J, Dwivedi R, Banjare P. Relying on Whom? Correlates of Out of Pocket Health Expenditure among the Rural Elderly in Odisha, India. Ageing International. 2017;42(3):306–23.

34. Gokhale SD. Towards a Policy for Aging in India. Journal of Aging & Social Policy. 2003;15(2-3):213–34.

35. Bharati K, Singh C. Ageing in India: need for a comprehensive policy. J IIM Bangalore Research Paper. 2013(421).

36. Dhillon P, Ladusingh L. Working life gain from gain in old age life expectancy in India. Demographic Research. 2013;28:733–62.

37. Ghosh S. Equity in the utilization of healthcare services in India: evidence from National Sample Survey. Int J Health Policy Manag. 2014;2(1):29–38.

38. Sun C. An analysis of RSBY enrolment patterns: Preliminary evidence and lessons from the early experience. India’s Health Insurance Scheme for the Poor: Evidence from the Early Experience of RSBY. New Delhi: Centre for Policy Research. 2011:84–116.

39. Narayana D. Review of the Rashtriya Swasthya Bima Yojana. Economic and Political Weekly. 2010;45(29):13–8.

40. Angell BJ, Prinja S, Gupt A, Jha V, Jan S. The Ayushman Bharat Pradhan Mantri Jan Arogya Yojana and the path to universal health coverage in India: Overcoming the challenges of stewardship and governance. PLOS Medicine. 2019;16(3):e1002759.

41. Brinda EM, Rajkumar AP, Enemark U, Prince M, Jacob KS. Nature and determinants of out-of-pocket health expenditure among older people in a rural Indian community. Int Psychogeriatr. 2012;24(10):1664–73.

42. Balarajan Y, Selvaraj S, Subramanian SV. Health care and equity in India. The Lancet. 2011;377(9764):505–15.

43. Anand A. Inpatient and outpatient health care utilization and expenditures among older adults aged 50 years and above in India. Health Prospect. 2016;15(2):11–9.

44. Carr E, Fleischmann M, Goldberg M, Kuh D, Murray ET, Stafford M, et al. Occupational and educational inequalities in exit from employment at older ages: evidence from seven prospective cohorts. Occup Environ Med. 2018;75(5):369–77.

45. Padula RS, Comper MLC, Moraes SA, Sabbagh C, Pagliato Junior W, Perracini MR. The work ability index and functional capacity among older workers. Brazilian Journal of Physical Therapy. 2013;17:382–91.

46. Silver MP, Dass AR, Laporte A. The effect of post-retirement employment on health. The Journal of the Economics of Ageing. 2018.

47. Buckley J, Tucker G, Hugo G, Wittert G, Adams RJ, Wilson DH. The Australian baby boomer population--factors influencing changes to health-related quality of life over time. J Aging Health. 2013;25(1):29–55.

48. d’Orsi E, Xavier AJ, Ramos LRJRdSP. Work, social support and leisure protect the elderly from functional loss: EPIDOSO study. 2011;45:685–92.

49. Li Y, Xu L, Chi I, Guo P. Participation in productive activities and health outcomes among older adults in urban China. Gerontologist. 2014;54(5):784–96.

50. Jang S-N, Cho S-I, Chang J, Boo K, Shin H-G, Lee H, et al. Employment status and depressive symptoms in Koreans: results from a baseline survey of the Korean Longitudinal Study of Aging. 2009;64(5):677–83.

51. Luo Y, Pan X, Zhang Z. Productive activities and cognitive decline among older adults in China: Evidence from the China Health and Retirement Longitudinal Study. Soc Sci Med. 2019;229:96–105.

52. Vives A, Gray N, Gonzalez F, Molina A. Gender and Ageing at Work in Chile: Employment, Working Conditions, Work-Life Balance and Health of Men and Women in an Ageing Workforce. Ann Work Expo Health. 2018;62(4):475–89.

53. Virtanen M, Lallukka T, Ervasti J, Rahkonen O, Lahelma E, Pentti J, et al. The joint contribution of cardiovascular disease and socioeconomic status to disability retirement: A register linkage study. Int J Cardiol. 2017;230:222–7.

54. McPhedran S. The labor of a lifetime?: health and occupation type as predictors of workforce exit among older Australians. J Aging Health. 2012;24(2):345–60.

55. Giles J, Wang D, Cai W. The labor supply and retirement behavior of China’s older workers and elderly in comparative perspective: The World Bank; 2011.

56. Majchrowska A, Broniatowska P. Ageing of the Workforce and Wages—Does the Type of Occupation Matter? Eastern European Economics. 2019;57(4):331–48.

57. Robroek SJ, Schuring M, Croezen S, Stattin M, Burdorf AJSjow, environment, health. Poor health, unhealthy behaviors, and unfavorable work characteristics influence pathways of exit from paid employment among older workers in Europe: a four year follow-up study. 2013:125–33.

58. Kim C-B, Yoon S-J, Ko J, editors. Economic Activity and Health Conditions in Adults Aged 65 Years and Older: Findings of the Korean National Longitudinal Study on Aging. Healthcare; 2017: Multidisciplinary Digital Publishing Institute.

59. Robroek SJ, Rongen A, Arts CH, Otten FW, Burdorf A, Schuring M. Educational Inequalities in Exit from Paid Employment among Dutch Workers: The Influence of Health, Lifestyle and Work. PLoS One. 2015;10(8):e0134867.

60. International Institute for Population Sciences (IIPS). Longitudinal Ageing Study in India (LASI) Wave 1, 2017-18, India Report. Mumbai: International Institute for Population Sciences; 2017–18.

61. NUEPA. EDUCATIONAL DEVELOPMENT INDEX. Department of Educational Management Information System National University of Educational Planning and Administration. 2009.

62. Cabral J, Vidaurre D, Marques P, Magalhães R, Silva Moreira P, Miguel Soares J, et al. Cognitive performance in healthy older adults relates to spontaneous switching between states of functional connectivity during rest. Scientific Reports. 2017;7(1):5135.

63. Singh P, Govil D, Kumar V, Kumar J. Cognitive Impairment and Quality of Life among Elderly in India. Applied Research in Quality of Life. 2017;12(4):963–79.

64. NSSO. Informal sector and conditions of employment in India. 2004.

65. NCO. National Classification of Occupations 2004 [Available from: https://labour.gov.in/sites/default/files/CodeStructure.pdf.

66. Tafuro S. An Economic Framework for Persisting Son Preference: Rethinking the Role of Intergenerational Support. Population Research and Policy Review. 2020;39(6):983–1007.

67. Adaji EE, Ahankari AS, Myles PR. An Investigation to Identify Potential Risk Factors Associated with Common Chronic Diseases Among the Older Population in India. Indian J Community Med. 2017;42(1):46–52.

68. Nilsson K, Hydbom AR, Rylander L. Factors influencing the decision to extend working life or retire. Scand J Work Environ Health. 2011;37(6):473–80.

69. Marengoni A, Angleman S, Melis R, Mangialasche F, Karp A, Garmen A, et al. Aging with multimorbidity: A systematic review of the literature. Ageing Research Reviews. 2011;10(4):430–9.

70. Cai L. The relationship between health and labour force participation: Evidence from a panel data simultaneous equation model. Labour Economics. 2010;17(1):77–90.

71. Aamir Ali S. Manual Scavenging: Intersection of Caste and Labor. International Journal of Law. 2019;5(5):2455–194.

72. Paltasingh T, Paliwal G. Tribal population in India: regional dimensions & imperatives. Journal of Regional Development and Planning. 2014;3(2):27–36.

73. Choobineh A, Shahnavaz H, Lahmi M. Major Health Risk Factors in Iranian Hand-Woven Carpet Industry. International Journal of Occupational Safety and Ergonomics. 2004;10(1):65–78.

74. Thongsuk W, Geater AF. Work-related discomfort among floor-sitting sedge weavers: a cross-sectional survey. International Journal of Occupational Safety and Ergonomics. 2021;27(2):523–34.

75. Singh AK, Meena ML, Chaudhary H, Karmakar S. Assessment of transmissibility of hand-arm vibration, noise exposure, and shift in hearing threshold among handicraft operatives’: a cross-sectional study. Journal of Industrial and Production Engineering. 2020;37(2-3):134–47.

76. Mahdavi N, Motamedzade M, Jamshidi AA, Darvishi E, Moghimbeygi A, Heidari Moghadam R. Upper trapezius fatigue in carpet weaving: the impact of a repetitive task cycle. International Journal of Occupational Safety and Ergonomics. 2018;24(1):41–51.

77. Kenny GP, Yardley JE, Martineau L, Jay O. Physical work capacity in older adults: Implications for the aging worker. American Journal of Industrial Medicine. 2008;51(8):610–25.

78. Robine J-M, Michel J-P. Looking Forward to a General Theory on Population Aging. The Journals of Gerontology: Series A. 2004;59(6):M590–M7.

79. Sampaio RF, Augusto VG. Aging and work: a challenge for the rehabilitation schedule. Brazilian Journal of Physical Therapy. 2012;16:94–101.

